# Long-term compositional stability of bacterial communities in frozen FMT capsules from a prospective donor cohort

**DOI:** 10.64898/2026.08.18.26360593

**Authors:** Elena Montenegro-Borbolla, Niklaus Johner, Keyvan Moser, Susanna Gerber, Maxime Audry, Aurélie Ballif, Carmen Chen, Benoît Guery, Claire Bertelli, Tatiana Galperine

## Abstract

**Background:** Faecal microbiota transplantation (FMT) is an effective treatment for recurrent *Clostridioides difficile* infections, yet the diversity of FMT formulations and delivery routes hampers comparisons across studies. Oral frozen capsules are widely used and current guidelines recommend storage at −80°C for up to two years. Despite extensive use of FMT, data on the long-term persistence and maintenance of their microbial composition remains limited.

**Method:** In this prospective study, we assessed the temporal stability of bacterial profiles in frozen FMT capsules derived from 48 donations of 10 healthy donors. Using metabarcoding, we longitudinally profiled one capsule per donation thawed within a month of production and after 3, 6, 12, and 24 months of storage. We used linear mixed effect models to evaluate changes in alpha diversity and community composition over time.

**Results:** Species richness remained stable across all timepoints, whilst species evenness decreased slightly. Although changes in community composition were detectable, they were small and mostly affected low-abundance genera. Clinical efficacy, assessed in a subset of recipients, was not associated with storage duration.

**Conclusion:** Our findings demonstrate that frozen FMT capsules preserve their bacterial community structure for at least two years of storage at −80°C, supporting their suitability for long-term biobanking and standardised clinical or research use.

## Introduction

Faecal microbiota transplantation (FMT) involves the transfer of processed stool from a healthy donor into the gastrointestinal tract of a recipient to restore microbial balance (Bascuñana et al., 2021; Nood et al., 2013). Its most established indication is the treatment of recurrent *Clostridioides difficile* infection (rCDI), a condition driven by antibiotic-induced disruption of the gut microbiota (Barbut and Meynard, 2002; Cheng et al., 2011; DePestel and Aronoff, 2013). In this context, FMT effectively re-establishes microbial diversity, addressing the underlying cause of recurrence where standard antibiotic therapies often fall short (Bascuñana et al., 2021; Minkoff et al., 2023). While current clinical guidelines primarily recommend FMT for rCDI (Johnson et al., 2021; Peery et al., 2024; Prehn et al., 2021), the use of FMT is increasingly investigated in a wide range of experimental applications, ranging from inflammatory bowel disease to metabolic syndrome and even neurological conditions (Cruz-Aguliar et al., 2019; Halkjær et al., 2023).

As its clinical use broadens, FMT has evolved significantly in terms of formulation and administration routes. Preparations now include suspensions and oral capsules (lyophilized or frozen), that can be administered via colonoscopy, nasogastric, nasoduodenal, or jejunal tube, as well as by enema or oral ingestion (Andary et al., 2024; Rakotonirina et al., 2022). Fresh preparations are no longer recommended in routine practice. The development of frozen formulations stored at −80°C with cryoprotective agents has significantly improved treatment feasibility and patient accessibility. Growing evidence suggests that the clinical effectiveness of FMT is generally comparable across different formulations, provided the microbial content is adequate (Gangwani et al., 2022; Shaheen et al., 2025). Nevertheless, the multiple alternatives for FMT formulations, pre-treatments, application routes, dosages, and even donor selection criteria complicate efforts toward standardisation (Aira et al., 2022; Kim et al., 2024; Rossier et al., 2023). Indeed, although donor screening follows general international guidelines, there is currently no universally accepted standard, and practices differ across countries. Among these challenges, ensuring the long-term stability of FMT products also remains a critical yet understudied consideration.

Several studies suggest that frozen FMT material can be stored for up to two years; however, the supporting evidence remains limited (Allegretti et al., 2020; Keller et al., 2021; Shaheen et al., 2025). Recent studies have begun to examine the stability of stored preparations in more depth, showing that frozen FMT capsules can maintain clinical efficacy rates above 75% even after one year of storage (Shaheen et al., 2025). The same study analysed longitudinally the bacterial composition of fresh and frozen products for a year and showed that dominant bacterial taxa were maintained across formulations, although their relative abundances differed (Shaheen et al., 2025). A decline in alpha diversity, measured by Shannon index, which considers both species richness and evenness, was observed over time, suggesting that alpha diversity may deteriorate during extended storage (Shaheen et al., 2025).

Building upon these findings, we investigated whether microbial diversity and clinical efficacy were maintained in FMT capsules produced and stored at −80°C for two years. After receiving market authorization from the national regulatory authority in 2024 (Swissmedic, marketing authorization no.: 68580), the CHUV FMT center is the sole facility in Switzerland accredited for the manufacturing of FMT treatments under stringent pharmaceutical standards. In addition, we explored transplant variability by analysing FMT material from different donors, as well as repeated donations from the same individuals across separate collection campaigns. Importantly, at CHUV FMT, each preparation is derived from a single donor, and materials from multiple donors are not pooled to produce one product. Overall, this study provides comprehensive long-term longitudinal data on the bacterial diversity and compositional stability of stored FMT preparations, while also evaluating how donor-related factors may influence the consistency and performance of these products.

## Materials and methods

### 1. FMT donor selection

FMT donors were recruited and screened according to the Swiss Agency for therapeutic products (Swissmedic) regulation based on the current version of the European Directorate for the Quality of Medicines & HealthCare (EDQM) at the CHUV FMT centre. In brief, donors are unpaid individuals between 18 and 50 years of age thoroughly screened to guarantee a safe FMT product, including questionnaires for personal and family history of diabetes, cancer, and immunological diseases, and for general health status; blood exams for normal physiological parameters; stool exams to detect enteric pathogens and multidrug-resistant bacteria, and SARS-CoV-2 test (detailed list on **Table S1**). Selected individuals donate stools twice a week for four weeks and are screened at the beginning and the end of this period, with annual follow-up for up to five years. If any of the exclusion criteria are met at either time point, the FMT material is discarded. All donors provided written informed consent for research purposes on the donation material.

### 2. FMT frozen capsules preparation and processing

FMT capsules were manufactured using material from a single donor under Good Manufacturing Practice (GMP) conditions at the CHUV Pharmacy. The FMT capsules followed the formulation described in reference (Youngster et al., 2014) and were stored at −80°C (BIOSTOOL, BB_041) (Huttner et al., 2019). All donations were performed on-site at CHUV and processed within six hours. Briefly, the production consisted of a filtered solution of donor faecal microbiota (50 g) homogenized with sterile saline (6 mL), filtered then centrifuged. After discarding the supernatant, the pellets were resuspended in glycerol, as a microbial cryopreserving agent, filtered and double-encapsulated in modified-release and ready-to-administer 0 and 00 capsules (Capsugel® DRCaps, Lonza, Switzerland).

Twelve donation campaigns between 2021 and 2023 from 10 different individuals were included in the study. Up to 5 capsules from each donation were stored at −80°C, separately from the medical products, and one capsule was thawed at each time point: during the first month after production (T0), and after 3-, 6-, 12-, and 24-month post-production. Samples from the same donation campaign were thawed and processed for DNA extraction at the same time to minimize technical biases.

The material of each capsule was extracted using a small needle and syringe, then transferred to a 15ml Falcon tube and diluted with 5ml of PBS. The mix was resuspended and transferred to a 2 ml screw-cap tube. Next, the 2 ml tubes were centrifuged at 15,000 g for 3 min and the supernatant was discarded. The pellet was resuspended in 500ml of PBS for DNA extraction on a MagNA Pure (Roche, Basel, Switzerland). Regions V3-V4 of the 16S rRNA gene were amplified for library preparation according to the protocol ‘16S Metagenomic Sequencing Library Preparation’ (Part. #15044223 Rev. B, Illumina, San Diego CA, USA) and reads were sequenced on Illumina MiSeq.

### 3. Bacterial community profiling and analysis

Raw reads were processed by zAMP, a pipeline based on DADA2 (Scherz et al., 2025), using the EzBioCloud as a taxonomic reference database, processed to contain only the V3-V4 region of the 16S genes. In total, this longitudinal study included 193 samples from 48 different donations. Seven samples were excluded, three due to insufficient sequencing depth and four due to the lack of longitudinal follow-up. Six more represented duplicate timepoints (same donation, same timepoint but different capsule) and were excluded from the main analysis after verifying that their composition was highly similar to the corresponding sample retained for the analysis (**Fig S1A**). Finally, eight samples were re-sequenced to investigate potential contamination during their processing as they displayed abnormal compositions and all originated from just 2 sequencing runs (out of 22). After comparing the re-sequenced data to the original data (**Fig S1 B**), we used the re-sequenced data for the main analyses presented in this paper. Note that the choices made above did not have any impact on the main results of the present study.

The remaining samples had an average sequencing depth of more than 450,000 reads per sample. To avoid biases due to sequencing depth we subsampled the reads for each sample to a sequencing depth yielding approximately 80,000 reads after the pre-processing steps of the pipeline (quality filtering, denoising, merging, chimera removal). The number of reads required for any given sample was determined by iteratively subsampling the raw reads, running the pre-processing steps of zAMP and then determining the number of reads to subsample in the next iteration. We obtained an average sequencing depth of 100,534 reads after sub-sampling, with considerable variability between samples (**Fig S2**), yielding an average of 80,011 ± 136 reads maintained after pre-processing.

All statistical analyses and data visualizations were performed using the R programming language version 4.2.3 (R-Core-Team, 2023). Alpha diversity metrics were calculated at species level using alpha() function from microbiome R package (Lahti and Shetty, 2019), selecting Chao1 index for richness and Pielou for evenness. To study the effect of storage time on alpha diversity, linear mixed effect (LME) models were fitted for each metric using the lme function from the nlme package (Pinheiro et al., 2023).

To assess the impact of long-term storage on microbiota composition, we performed a Constrained Analysis of Principal Coordinates using the *capscale* function from the *vegan* package (Oksanen et al., 2001). The analysis was based on a Bray-Curtis dissimilarity matrix computed from relative abundance data, filtered for a minimum prevalence of 10%, and was used to quantify compositional variation in microbial communities while controlling for experimental design.

To identify the main taxa changing with storage time, we used the *MaAsLin2* package. Genus-level counts per sample were normalised with total sum scaling (TSS), LOG transformed and filtered with a 10% minimum prevalence cutoff using the *MaAsLin2* function options. Significantly differentially abundant genera, after false discovery rate (FDR) correction (q-val <0.05), were plotted using the *ggplot2* package (Wickham, 2016).

For all statistical models described above, storage time (in months) was included as a fixed effect, while donor, campaign, and donation (nested within donor and campaign) were modelled as random effects to account for the longitudinal and hierarchical structure of the data.

### 4. Effect of storage time on clinical efficacy of FMT capsules

Eligible patients who received FMT as part of routine clinical care for rCDI were prospectively included, along with their corresponding donors involved in the production of the FMT batches used for treatment. Ethical approval for this study was obtained from the Canton of Vaud Ethics Committee (CER-VD 2018-01330, AO-2025-00084). Written informed consent was obtained for donors for participation in the research and reuse of biological samples. A multivariable logistic regression model was used to examine the association between storage time of the FMT product and clinical success. Storage time (in days), patient age (in years), and sex (male/female) were included as independent variables. Odds ratios (ORs) and 95% confidence intervals (CIs) were reported.

## Results

Storage of FMT capsules at −80°C slightly reduces species evenness, with a non-significant trend in richness.

We modelled Chao1 richness as a function of storage time using a linear mixed-effects model with random intercepts for donor, campaign, and donation. The fixed effect of storage time (months) was not statistically significant (β = −0.55 ± 0.34, t(131) = −1.62, p = 0.11), indicating a slight but non-significant decrease in richness over time (**Figure 1A**). Most of the variance occurred between donors (SD = 33.27) and between campaigns (residual SD = 18.76), with negligible contribution from donations. Subsequently, we examined species evenness by modelling the Pielou index as a function of storage time, using the same approach. In contrast to richness, the fixed effect of storage time was statistically significant (β = −0.0008 ± 0.0002, t(131) = −3.52, p < 0.001), indicating a small but consistent decrease in species evenness over time (**Figure 1B**). Here, the majority of variance was observed between donors (SD = 0.03), followed by donations (residual SD = 0.019), and campaigns (residual SD = 0.009).

**Figure 1:**
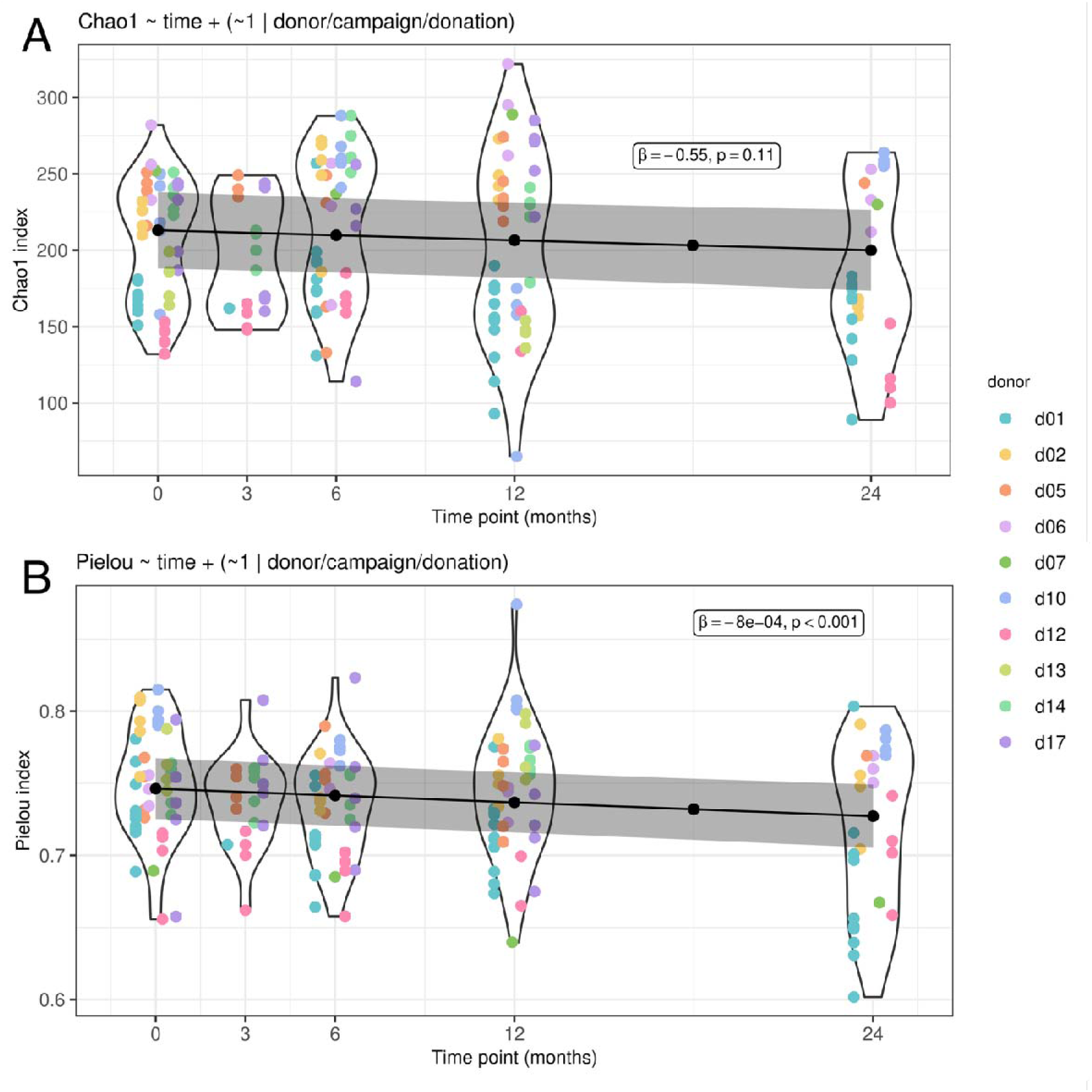
Alpha diversity metrics at species level across time of capsules storage at −80°C. The regression line shows the fixed effect of time from a linear mixed-effects model with random intercepts for donor, campaign, and donation, for both the Chao1 richness index (**A**) and the Pielou evenness index (**B**). Although richness decreased slightly over time (β = −0.55 ± 0.34), this trend was not statistically significant (t(131) = −1.62, p = 0.11). A similar trend is observed for evenness, which slightly decreases over time (β = −0.0008 ± 0.0002), although this trend is statistically significant (t(131) = −3.52, p<0.001). Colours represent the different donors.

Changes in community structure evidence small decrease in *Bacteroides, Alistipes* and other rare genera.

A Canonical Analysis of Principal Coordinates with storage time as constrained variable and donor, campaign, and donation as conditional variables returned a statistically significant model (p-value < 0.001, **Fig 2A**). These variables combined explained 86% of variance, of which only 0.8% was attributed to storage time. This demonstrates that storage at −80°C over a two-year period has a minimal impact on bacterial composition. An analysis of differentially abundant genera over storage time identified 4 genera whose change in abundance may contribute to the slight compositional shift observed in the bacterial communities (**Fig 2B**). Most of these genera are present at low relative abundance (below 0.1%) across the majority of samples, with the notable exception of *Bacteroides*, which constitutes up to 50% of the community in some cases (**Fig 2B, Fig S3, Fig S4**). *Sutterella* reaches a relative abundance of 3% in certain samples, whereas *Pumilibacter* and PAC001360 (a bacterium belonging to the *Borkfalkiaceae* family) are generally rare. All genera presenting a significant differential abundance exhibited decreasing trends over time, with slope estimates ranging from −0.053 to −0.021. Overall, the magnitude of these changes is small and does not affect the global compositional structure, suggesting a minor relevance of storage to potential biological effects.

**Figure 2:**
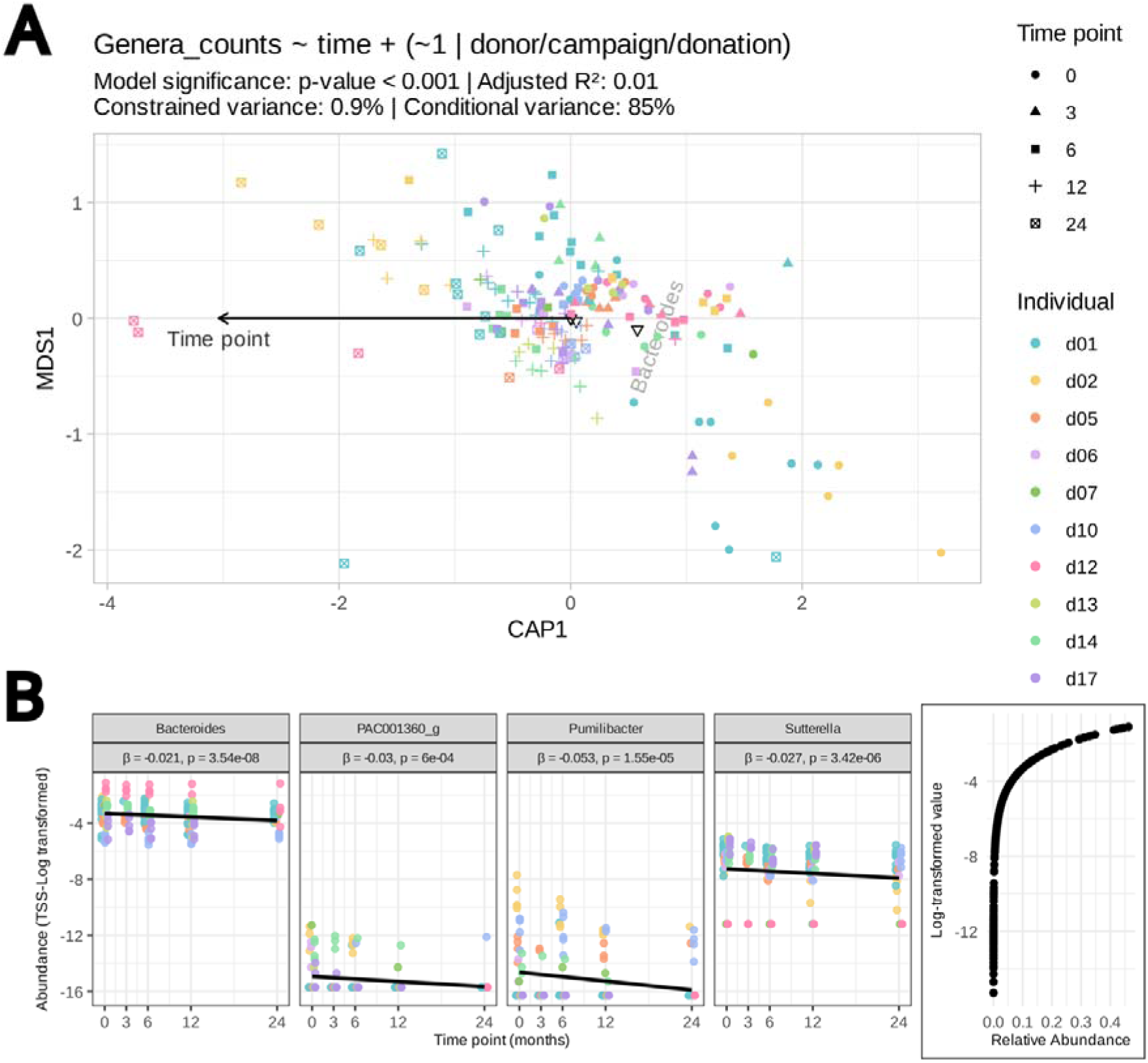
Compositional changes with FMT capsules storage time at −80°C. **A)** Canonical Analysis of Principal Coordinates (CAP) with the function capscale() on the Bray-Curtis distance at the genus level. Model statistics and explanatory power are indicated below the model function. **B)** Genera associated with the storage time identified by MaAsLin2 with an adjusted p-value < 0.05. The y-axis represents the TSS-log transformed data, and the bottom right panel shows the relation between the relative abundance and the transformed values to facilitate the interpretation of the data. Colour coding represents donors.

Clinical efficacy of the FMT materials used.

To evaluate the efficacy of the FMT capsules after long-term storage, we retrospectively analysed whether storage duration at −80°C was associated with a change in clinical efficacy using a multivariable logistic regression model. This analysis included 56 FMT administrations derived from 29 stool donations obtained from the 8 donors included in this study delivered to 56 recipients. Overall, clinical success was achieved in 91.1% (51/56) of recipients. We found no association between clinical success and storage time (**Table 1**). Patient sex and age were also incorporated in the model, but neither yielded a significant result (**Table 1**).

**Table 1:** Odds ratio for the success of frozen FMT capsules using a multivariable logistic regression model. Total number of samples = 56.

| Variable | Values | Odds Ratio | 95% CI | p-value |
| --- | --- | --- | --- | --- |
| Storage time (days) | 444 (201)<sup>1</sup> | 1.00 | 0.99 – 1.00 | 0.5 |
| Age (years) | 62 (20)<sup>1</sup> | 0.97 | 0.90 – 1.02 | 0.3 |
| Sex (male) | M=17<br>F=39 | 1.67 | 0.21 – 34.8 | 0.7 |

<sup>1</sup>Mean (standard deviation). CI = Confidence interval

## Discussion

In this study, we assessed the impact of long-term storage on the metagenomic microbial composition of FMT capsules produced at the sole FMT manufacturing site in Switzerland. Capsules were generated from 48 stool donations obtained from 10 individual donors included in this study, stored at - 80°C, and analysed longitudinally over a two-year period. Using linear mixed-effects models, we observed minimal changes in both alpha and beta diversities, with only a slight reduction in species evenness. This modest shift was accompanied by a small decrease in the relative abundance of the *Bacteroides* genus, along with minor changes in rare taxa. Consistent with these microbiological findings, an evaluation of clinical outcomes from 29 of the stored donations originating from the same donor cohort, which were used for 56 recipients, showed no association between storage duration and clinical efficacy.

Frozen FMT capsules have revolutionised FMT by allowing banking and on-demand use, but it is critical to better characterise stability over time. Previous studies showed that frozen stools retain clinical effectiveness for at least 6-12 months (Facchin et al., 2025; Shaheen et al., 2025). However, data beyond one year are sparse and earlier guidelines were based more on expert opinion than on evidence (Shaheen et al., 2025). While the exact mechanisms underlying the efficacy of FMT remain unclear (Ianiro et al., 2022; Schmidt et al., 2022), the bacterial component of FMT has been extensively studied, and restoration of microbial diversity has emerged as a key hallmark of successful treatment. Accordingly, assessing the stability of bacterial diversity and community structure over extended storage periods represents a biologically relevant and pragmatic approach to evaluating long-term product integrity when paired with an analysis of clinical treatment efficacy. By longitudinally examining capsules stored for up to 24 months, we aimed to determine whether prolonged storage alters the bacterial community structure in ways that could potentially influence efficacy and, ultimately, ensure consistent product quality for patients.

In this study, we evaluated the alpha diversity and compositional changes of frozen FMT capsules over a two-year storage period. To our knowledge, this represents the largest dataset specifically dedicated to this purpose. A recent study by Facchin and colleagues reported a similar analysis based on eight samples from two FMT donors, observing comparable alpha diversity (Shannon index) between fresh samples and samples stored for 24 months at −80°C in 10% glycerol (Facchin et al., 2025). While informative, the limited number of samples and donors in that study constrains the ability to account for inter-donor variability and potential confounding factors and reduces statistical power to detect subtle storage-related effects. In contrast, our larger and longitudinally structured dataset enables a more robust assessment of diversity trends over time.

Another study, which compared the clinical effectiveness of lyophilised versus frozen FMT products and assessed the impact of storage duration up to 12 months, concluded that storage time led to a decrease in bacterial diversity as measured by the Shannon index (Shaheen et al., 2025), which reflects both species richness and evenness. This is consistent with our results showing slight decreases in both richness and evenness over time, confirming that prolonged freezing may lead to a modest reduction in alpha-diversity.

Encouragingly, a separate study using RNAlater to preserve stool samples stored at −80°C for five years reported minimal effects on the integrity of the gut microbiota. While RNAlater is not compatible with FMT applications due to cell lysis, this work provides a useful reference for the extent of microbiota variation that can arise from long-term storage and downstream processing alone. In that study, minor changes in the relative abundance of certain genera were correlated with the initial microbial composition and remained smaller than both inter-individual variability and variability between sequencing runs (Tap et al., 2019). In line with this, our analysis revealed subtle but measurable shifts in the bacterial community over time, predominantly affecting genera with low relative abundance, and of a magnitude consistent with expected technical and analytical variability rather than biologically meaningful compositional change.

Freezing after FMT production can affect bacterial groups differently. In our dataset, we observed a slight decrease in the abundance of the *Bacteroides* genus, consistent with findings from the aforementioned preservation study using RNAlater (Tap et al., 2019), and another study evaluating FMT suspensions stored frozen for 18 months (Dorsaz et al., 2020). In the latter study, the authors differentiated between intracellular DNA and extracellular DNA, including DNA from human cells and damaged bacterial cells, which revealed that Gram-negative anaerobes are particularly sensitive to prolonged freezing. In contrast, more resilient taxa predominantly belonged to the phylum *Bacillota*, suggesting that features such as thicker cell walls and the ability to form spores confer increased resistance to both the manufacturing process and long-term frozen storage (Dorsaz et al., 2020; Shaheen et al., 2025). These observations align with our findings: *Bacillota* was the most abundant phylum in our samples (**Error! Reference source not found.B**), and none of the taxa identified as differentially abundant were known spore-formers (Carro and Trujillo, 2015; Hiippala et al., 2020, 2016; Morotomi et al., 2012; Parker et al., 2020; Wexler, 2007; Zoetendal et al., 2003).

In this study, we focused on bacterial DNA and on relative abundance metrics; thus, we did not distinguish between viable and non-viable fractions (Shaheen et al., 2025) or between intracellular and extracellular or degraded bacterial DNA (Dorsaz et al., 2020). Incorporating these additional layers of analysis could provide a more detailed understanding of functional microbial preservation during extended storage.

Previous work supports the notion that modest storage-related compositional changes do not necessarily compromise the clinical efficacy of FMT products. In a recent study, Facchin and colleagues reported that frozen FMT preparations retained clinical effectiveness despite minor microbiota shifts associated with prolonged storage, suggesting a degree of functional resilience of the bacterial community (Facchin et al., 2025). In contrast, another study evaluating clinical outcomes across different storage durations identified storage time as a potential factor influencing efficacy in specific subgroups, particularly among elderly recipients, highlighting the possibility of increased susceptibility to subtle product variations in vulnerable populations (Shaheen et al., 2025). In our cohort, however, we observed no association between clinical efficacy and storage duration, nor with patient age or sex, suggesting that within the storage conditions and population studied, prolonged freezing did not measurably impact treatment success. Small sample size (n=56) and, more importantly, the low number of recurrences (n=5, 8.9%) limited the power of the multivariate logistic regression, which included three covariates. The wide confidence intervals seen for all odds ratios are likely a reflection of this limited power rather than a true absence of association.

Our results demonstrating the long-term stability of frozen FMT capsules have important implications for expanding FMT accessibility. In addition to the preservation of microbial composition for up to two years, we observed no evidence that prolonged storage adversely affected clinical efficacy, reinforcing the functional robustness of these products. Together, these findings support the feasibility of maintaining larger stool bank inventories, which could improve patient access by reducing reliance on frequent fresh donations. Nevertheless, implementing long-term monitoring protocols remains needed to detect potential degradation before product distribution and to ensure sustained product quality over time.

## Conclusion

Frozen FMT capsules stored at −80°C exhibited measurable yet minimal compositional changes over a two-year period, as observed using 16S rRNA gene sequencing, without any measurable change in efficacy of the FMT in our cohort. The demonstrated stability of their bacterial composition strengthens confidence in the long-term storage of frozen FMT products and suggests that extended biobanking does not compromise clinical efficacy. As FMT continues to expand into therapeutic areas beyond recurrent *C. difficile* infection, including inflammatory, metabolic and even neurological disorders, ensuring product consistency will be essential for generating reliable clinical outcomes. Our findings therefore provide a foundation for harmonising FMT preparation and storage practices across both research and clinical settings.

## Acknowledgments

We thank the Laboratory of Genomics and Metagenomics and the sequencing platform of the DMLP (CHUV) for their technical assistance.

## Availability of data and materials

The raw sequencing data generated in this study have been deposited in the European Bioinformatics Institute (EBI) repository and are available at https://www.ebi.ac.uk/ena/browser/view/PRJEB107981. The analysis scripts used to process and analyze the data are publicly available at https://doi.org/10.5281/zenodo.21371874.

## Author contributions

Conceptualization, Methodology, Investigation, Formal analysis, Data curation: TG, BG, EM, NJ, KM, MA, AB, CC. Writing – Original Draft: EM, BG. Writing – Review & Editing: EM, NJ, SG, CB, TG, BG. Supervision: TG, BG, CB. Funding Acquisition: TG, BG, CB.

## Ethical approval and consent to participate

Ethical approval for this study was obtained from the Canton of Vaud Ethics Committee (CER-VD 2018-01330, AO-2025-00084). Written informed consent was obtained for donors for participation in the research and reuse of biological samples.

## Declaration of AI and AI-assisted technologies in the writing process

We acknowledge the use of Claude (Anthropic) for language refinement in the preparation of this manuscript. After using this tool, the authors reviewed and edited the content as needed and take full responsibility for the content of the publication.

