## Supplemental material for "Long-term compositional stability of bacterial communities in frozen FMT capsules from a prospective donor cohort"

**Supplementary Material**

**Table S1:** Biological screening for donor selection in fecal microbiota transplantation for the treatment of Clostridioides difficile infection

| Blood Analysis | |
| --- | --- |
| Haematology | full blood count |
| Chemistry | Creatinine, C reactive protein, albumin, liver enzymes, glycated hemoglobin |
| Viruses | Hepatitis A (IgM anti-HAV), B (HBsAg, total anti-HBc, anti-HBs), Hepatitis C antibody, and hepatitis E (IgM, IgG), HIV (anti-HIV 1/2, p24 antigen), HTLV-1/2 (anti-HTLV-1/2) |
| Bacteria | *Treponema pallidum* antibodies (TPHA) |
| Parasites | *Strongyloides stercoralis* (serology) (only if travel to a risk area), *Entamoeba histolytica* (serology) |
| Stool Analysis | |
| Bacteria | *Clostridioides difficile*, *Campylobacter jejuni/coli*, *Salmonella spp*, *Shigella spp*, Entero-invasive *E. coli*, shiga toxins STx1 and STx2, *Yersinia enterocolitica*, *Helicobacter pylori* |
| Viruses | Adenovirus, rotavirus, norovirus groups I and II, *Hepatitis E*, SARS-CoV-2 |
| Parasites | *Giardia intestinalis*, *Cryptosporidium hominis/parvum*, *Entamoeba histolytica*, helminths, *Dientamoeba fragilis*, *Strongyloides stercoralis*, *Blastocystis hominis* |
| Multi-resistant bacteria: | carbapenemase-producing Enterobacterales, extended-spectrum beta-lactamases vancomycin-resistant enterococci, Methicillin-resistant *Staphylococcus aureus* |
| Immunological marker | Calprotectin |
| For severely immunosuppressed patients | |
| Blood | Cytomegalovirus (CMV) IgM and IgG |
| Viruses | Epstein-Barr virus (EBV) IgM and IgG, toxoplasmosis (IgG, IgM) |
| Stool | *Plesiomonas shigelloides*, parechovirus, astrovirus, enterovirus, sapovirus, *Cyclospora*, *Isospora*, *Microsporidia* |

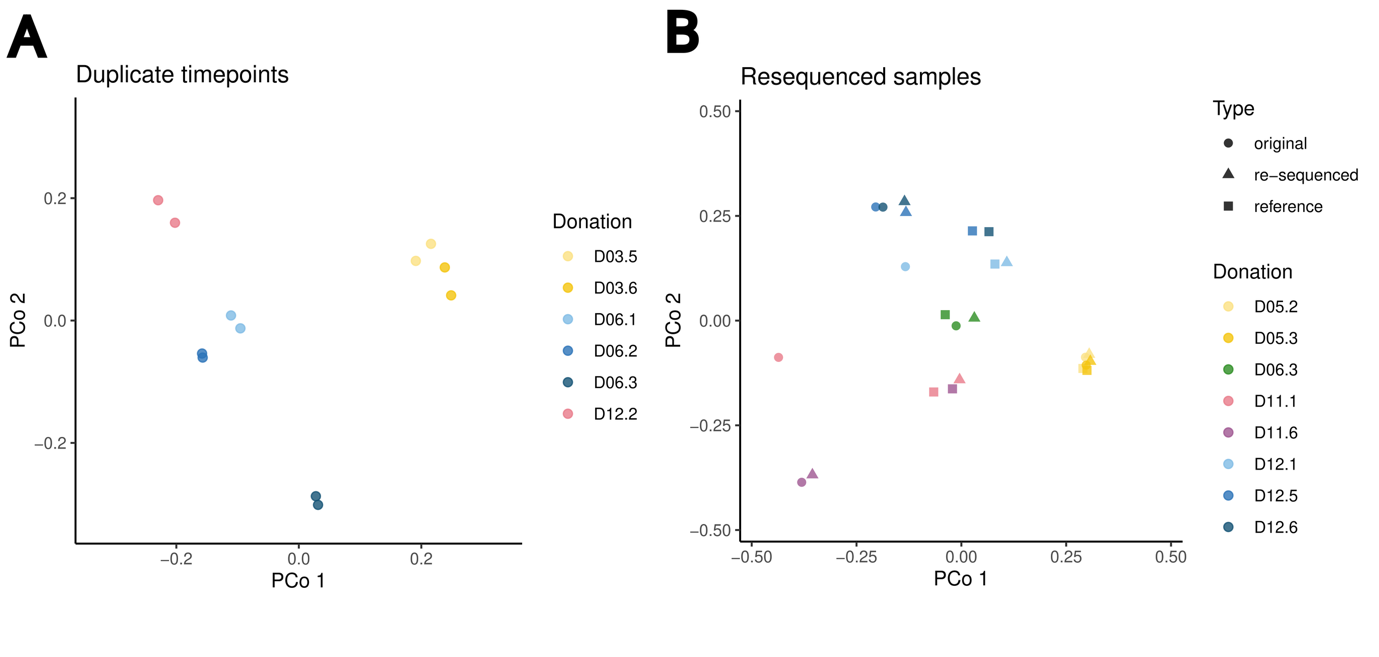

**Figure S1:** Ordination plots showing the first two principal coordinates from the Bray-Curtis dissimilarities for (A) donations with two different samples for the same timepoint, and (B) donations which were re-sequenced due to suspected contaminations, with circles representing the original data, triangles the re-sequenced data and the squares are reference samples from timepoint T0 for the same donations.

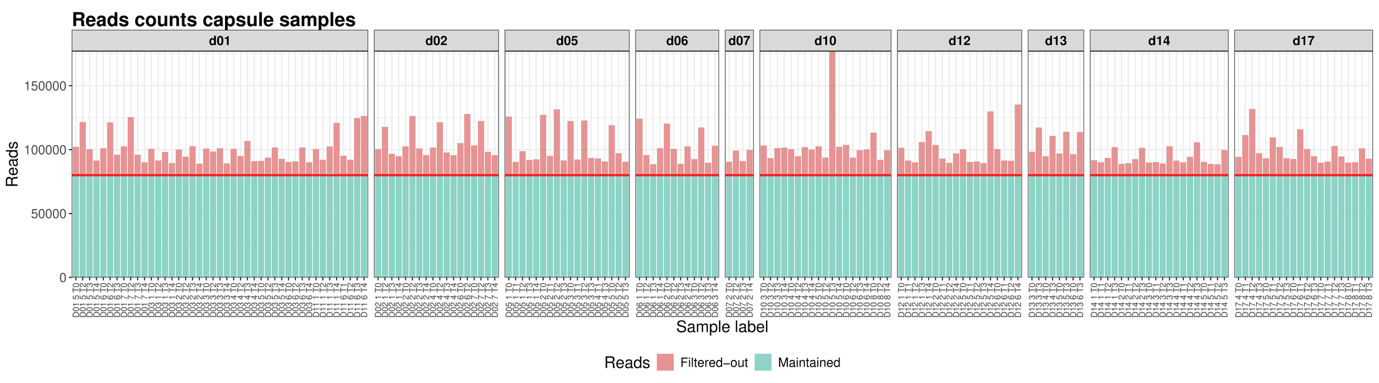

**Figure S2:**Number of reads maintained after pre-processing. Height of the bars corresponds to the number of reads after subsampling, in red the number of reads lost during the pre-processing steps and in blue the number of reads remaining after preprocessing. Horizontal red line represents the target value of 80,000 maintained reads.

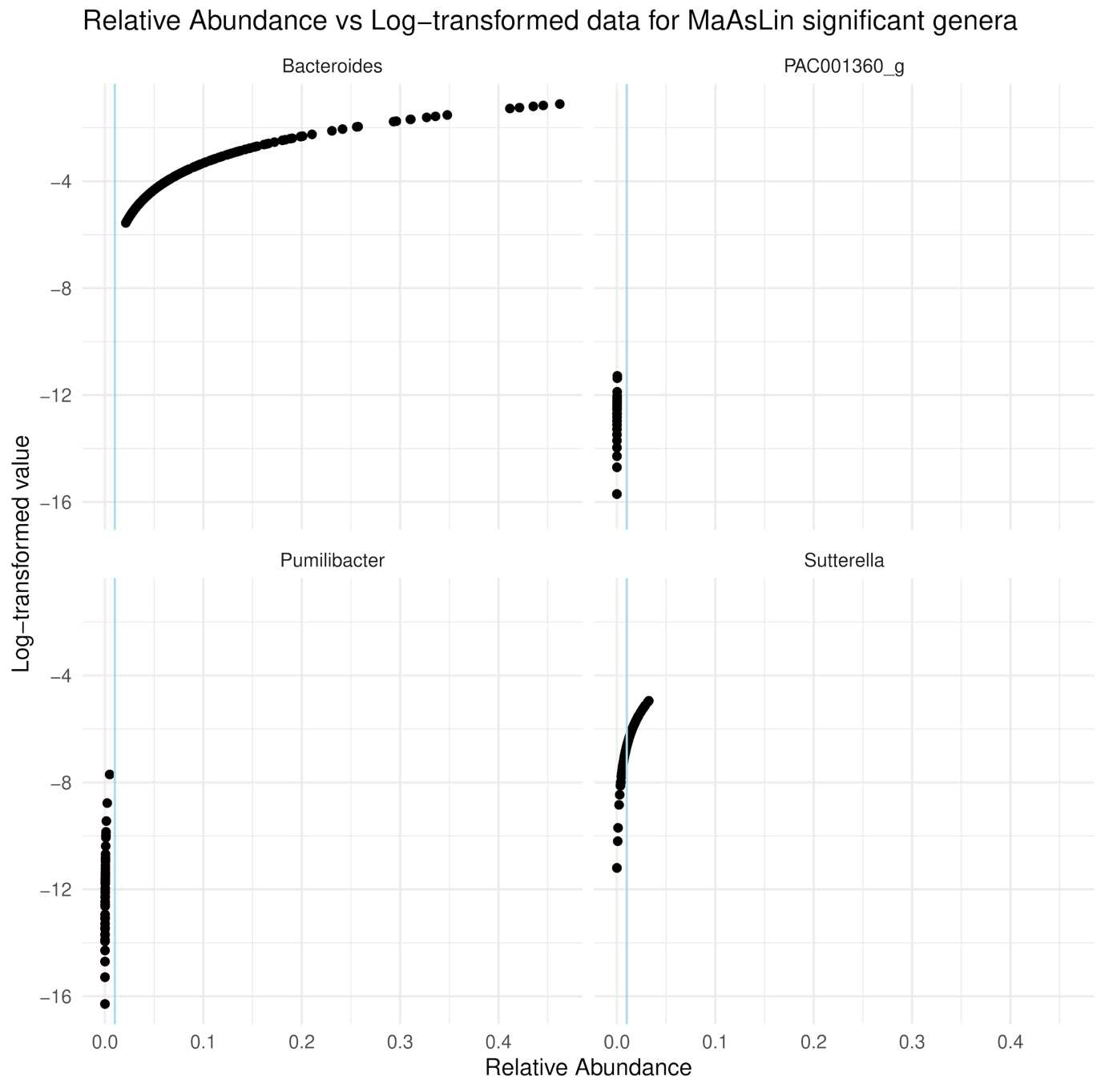

**Figure S3:** Relation between relative abundance (range 0-1) and CLR-transformed abundances of the genera of interest identified by MaAsLin2, intended to facilitate species abundance interpretation. Dots represent the sample counts of each genus in each sample. Vertical light blue line designates 0.01 (1%) relative abundance.

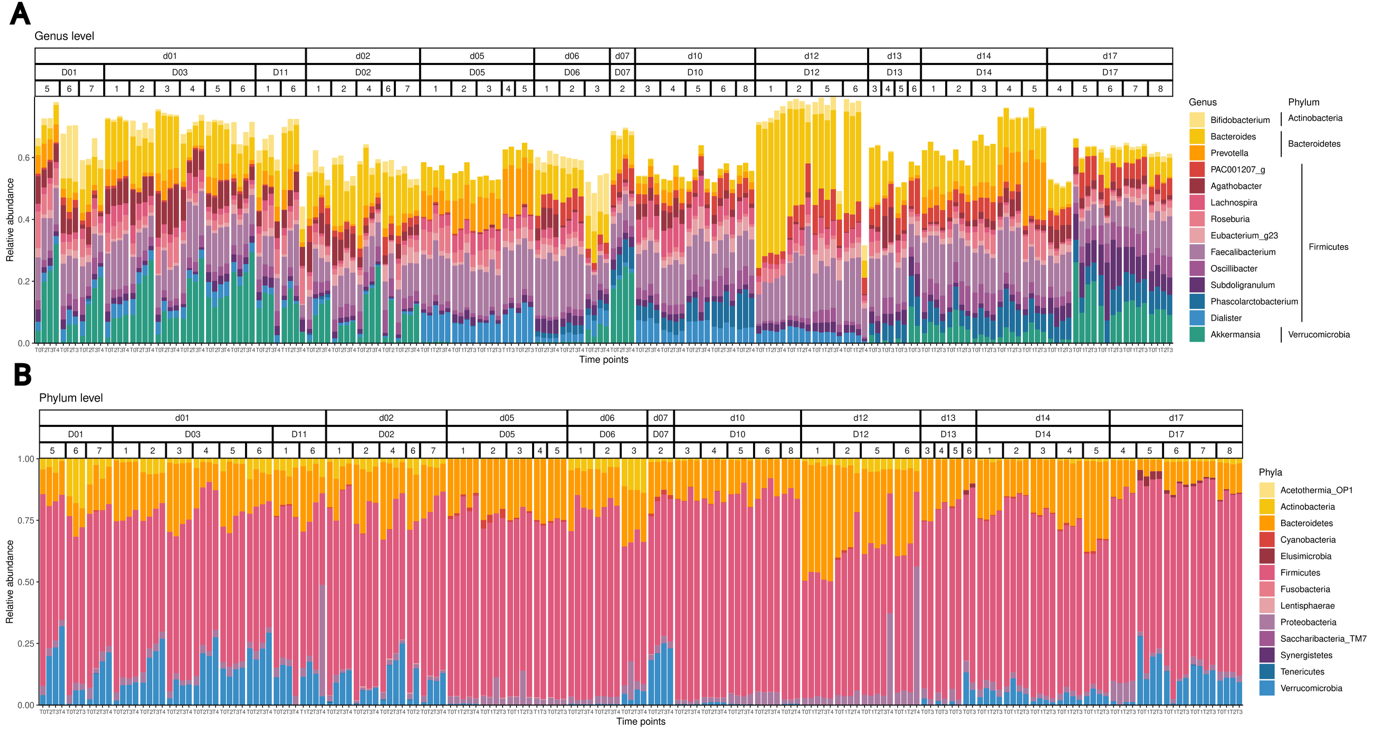

**Figure S4**: Relative abundance (range 0-1) of the 14 most abundant **genera (A)** and **phyla (B)**, grouped by donor, donation campaign, and donation number. Empty space on (A) reflects other taxa. Note that Firmicutes correspond to Bacillota in newer taxonomies.
